# Pathway-specific genomic alterations in pancreatic cancer across diverse cohorts

**DOI:** 10.1101/2025.02.27.25323058

**Authors:** Cecilia Monge, Brigette Waldrup, Francisco G. Carranza, Enrique Velazquez-Villarreal

## Abstract

**Background/Objectives:** Pancreatic cancer (PC) is an aggressive malignancy with rising incidence and poor survival rates. While Hispanic/Latino (H/L) patients have a lower overall incidence compared to Non-Hispanic White (NHW) patients, they are diagnosed at younger ages, often present with more advanced disease, and experience worse survival outcomes. The molecular drivers underlying these disparities remain poorly understood. Key oncogenic pathways, including TP53, WNT, PI3K, TGF-Beta, and RTK/RAS, play crucial roles in tumor progression, therapy resistance, and response to targeted treatments. However, their ethnicity-specific alterations and prognostic implications in PC remain largely unexplored. This study aims to characterize pathway-specific mutations in PC among H/L and NHW patients, assess tumor mutation burden, and identify ethnicity-specific oncogenic drivers using publicly available datasets. The findings may provide critical insights to optimize precision medicine strategies and enhance targeted therapies for underrepresented populations.

**Methods:** A bioinformatics analysis was performed using publicly available PC datasets to evaluate mutation frequencies in genes associated with the TGF-Beta, RTK/RAS, WNT, PI3K, and TP53 pathways. The study included 4,248 patients, with 407 identified as H/L and 3,841 as NHW. Patients were stratified by ethnicity to assess differences in mutation prevalence. Chi-squared tests were conducted to compare mutation rates between groups, while Kaplan-Meier survival analysis was performed to evaluate overall survival differences based on pathway-specific alterations.

**Results:** Significant differences were observed in the TGF-Beta pathway between H/L and NHW patients. TGF-Beta mutations were less prevalent in H/L patients (18.4% vs. 24.4%, p = 8.6e-3). Additionally, genes related to the TGF-Beta pathway showed significant alterations, with SMAD2 (1.5% vs. 0.4%, p = 6.3e-3) and SMAD4 (15% vs. 19.9%, p = 0.02) exhibiting notable differences. Although RTK/RAS, WNT, PI3K, and TP53 pathway mutations were not statistically significant overall, borderline significance was observed in genes associated with these pathways, including ERBB4 (3.4% vs. 1.8%, p = 0.03), ALK (2.7% vs. 1.1%, p = 0.01), HRAS (1.2% vs. 0.1%, p = 1.3e-4), and RIT1 (0.7% vs. 0.1%, p = 0.03) in the RTK/RAS pathway, as well as CTNNB1 (2.9% vs. 1.3%, p = 0.01) in the WNT pathway. Survival analysis revealed no significant differences in overall survival among H/L patients. However, NHW patients with TP53 pathway alterations exhibited borderline significant differences in survival outcomes.

## 1. Introduction

Pancreatic cancer (PC) is an aggressive malignancy and one of the leading causes of cancer-related mortality worldwide, with incidence rates continuing to rise (1,2). Despite advances in treatment, PC remains associated with poor survival outcomes due to its late-stage diagnosis, aggressive tumor biology, and high resistance to conventional therapies (3,4). The disease is often asymptomatic in its early stages, leading to delayed detection and limited treatment options, contributing to one of the lowest five-year survival rates among all cancers (5). Given its increasing public health burden, understanding the genomic and clinical factors influencing PC progression is essential for improving risk prediction, early detection, and therapeutic strategies.

While Hispanic/Latino (H/L) individuals have a lower overall incidence of PC compared to Non-Hispanic White (NHW) patients, they are diagnosed at younger ages and at more advanced disease stages, leading to significantly poorer survival outcomes (6). Additionally, the high prevalence of diabetes mellitus in the H/L population—a known risk factor for PC—has been linked to an increased likelihood of developing the disease (7,8). However, genomic profiling of PC in H/L patients has been historically limited, creating a critical gap in understanding ethnicity-specific molecular drivers of disease progression and treatment response. Prior studies have identified actionable somatic mutations in PC that may inform precision medicine approaches, but the extent to which these mutations differ between racial/ethnic groups remains unclear (9,10).

PC is characterized by frequent alterations in key oncogenic pathways, including TGF-Beta, RTK/RAS, WNT, PI3K, and TP53, which regulate essential cellular functions such as proliferation, apoptosis, DNA repair, and immune evasion (11,12). Dysregulation of these pathways is associated with tumor aggressiveness, resistance to chemotherapy, and poor clinical outcomes. However, the impact of these molecular alterations within H/L populations remains largely unexplored.

The TGF-Beta signaling pathway has a dual role in PC, acting as a tumor suppressor in early stages but promoting epithelial-to-mesenchymal transition (EMT), invasion, and metastasis in advanced disease (13). In PC, mutations in SMAD4 and TGFBR2 are commonly observed and have been implicated in disease progression and therapy resistance (14). However, the frequency and functional consequences of these alterations in H/L patients require further investigation.

The RTK/RAS pathway plays a central role in PC pathogenesis, with KRAS mutations occurring in over 90% of cases, driving uncontrolled cell growth and conferring resistance to targeted therapies (15,16). Mutations in additional RTK/RAS pathway genes, including ERBB4, ALK, and HRAS, may further contribute to tumor progression and therapeutic resistance (17,18). Given the high prevalence of KRAS-driven tumors in PC, understanding ethnicity-specific variations in this pathway is crucial for developing personalized treatment strategies.

The PI3K/AKT pathway is a critical regulator of tumor metabolism and survival, with frequent alterations in PTEN and PIK3CA driving increased tumor invasiveness and immune evasion (19). Mutations in PI3K pathway genes have been associated with poor prognosis and treatment resistance, making them important targets for precision medicine approaches (20,21).

The TP53 pathway plays a fundamental role in genomic stability, apoptosis, and cell cycle regulation. TP53 mutations are among the most common alterations in PC, and emerging evidence suggests that mutant p53 interacts with KRAS to drive metastasis (23,24). In addition to TP53 mutations, CDKN2A deletions are frequently observed in PC and are associated with aggressive tumor behavior (24,26). Ethnicity-specific differences in TP53 pathway alterations could have significant implications for PC risk stratification and treatment response.

The WNT/β-catenin signaling pathway is another key driver of PC progression, regulating cell proliferation and differentiation. WNT pathway mutations have been identified in nearly all PC cases, with alterations in genes such as CTNNB1 and RNF43 contributing to tumor initiation and progression (27). Aberrant WNT signaling has also been linked to chemoresistance, underscoring the need for pathway-specific therapeutic interventions (28).

Given the rising incidence of PC and the disproportionately poor outcomes observed in H/L patients, a comprehensive genomic analysis of these key oncogenic pathways is critical for advancing risk prediction, precision medicine, and targeted therapeutic strategies, as has been suggested for other cancers (29–35). This study aims to characterize pathway-specific mutations in TP53, WNT, PI3K, TGF-Beta, and RTK/RAS signaling among H/L and NHW patients, assess tumor mutation burden, and evaluate the prognostic implications of these alterations. By identifying ethnicity-associated molecular differences, this study seeks to inform clinical epidemiology and risk prediction models, ultimately improving early detection and personalized treatment approaches for PC in underrepresented populations.

## 2. Materials and Methods

For this analysis, we leveraged clinical and genomic data from 14 PC datasets accessed via the cBioPortal database. These datasets included studies categorized under PC, as well as data from the GENIE Cohort v17.0-public dataset. To refine our sample pool, we applied strict inclusion criteria, selecting only patients identified as H/L or NHW. Following dataset selection and filtering, four datasets met all criteria, comprising 407 H/L PC patients. For comparison, 3,841 NHW PC patients were included using identical criteria (Tables 1 & 2). This study represents one of the most comprehensive investigations of TP53, WNT, PI3K, TGF-Beta, and RTK/RAS pathway alterations in an underrepresented population, offering key insights into molecular disparities in PC.

Patients were stratified based on ethnicity (H/L vs. NHW) and further categorized according to the presence or absence of TP53, WNT, PI3K, TGF-Beta, and RTK/RAS pathway alterations. This classification allowed a detailed examination of the interactions between genetic alterations and ethnicity. Table 1 presents the number of patients included in the analysis, providing a breakdown of cases by racial/ethnic group. By comparing mutation prevalence between H/L and NHW patients, this study aims to characterize the molecular differences in PC and their implications for precision medicine and targeted therapies.

**Table 1.**
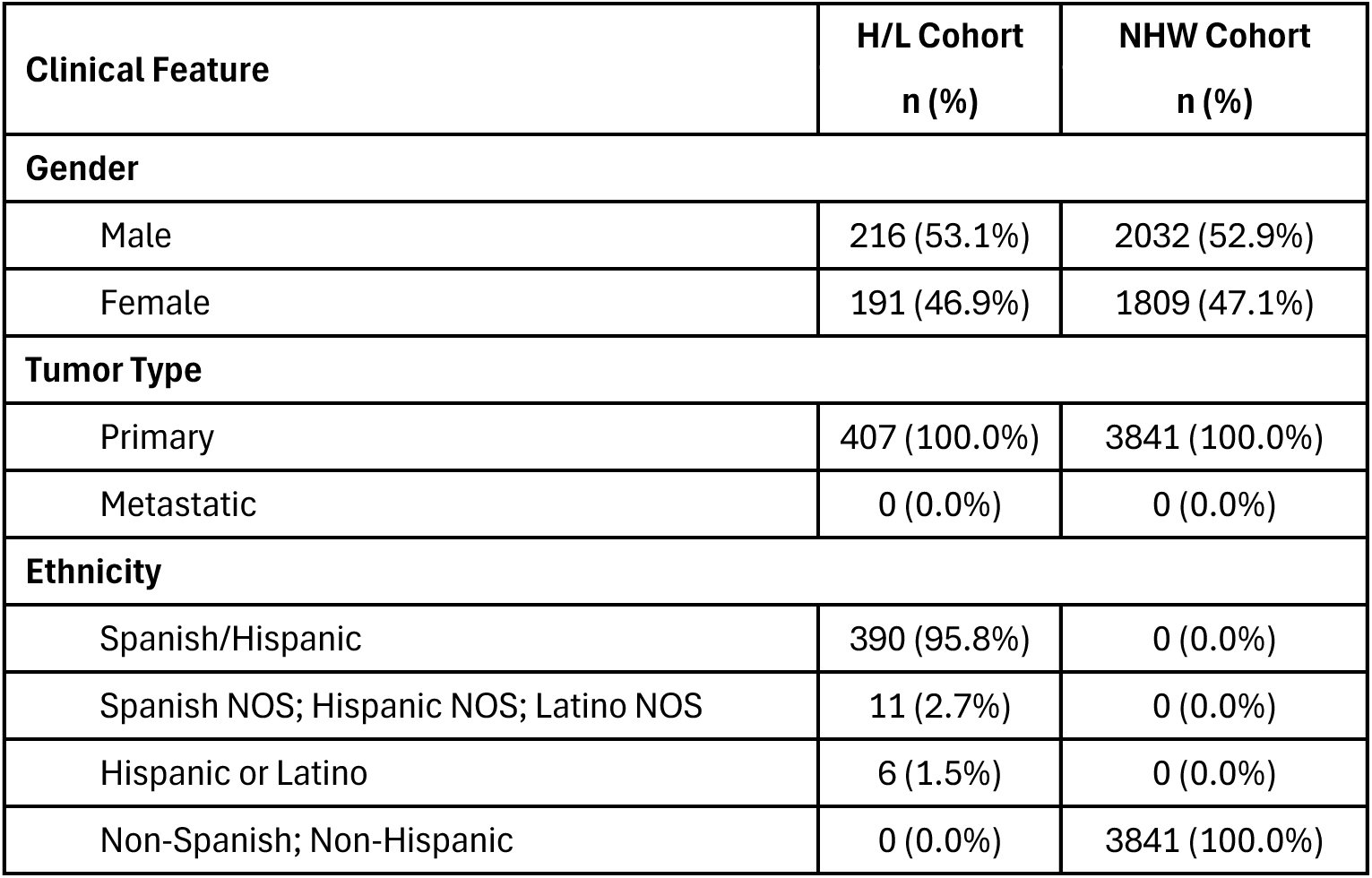
Patient Demographics and Clinical Characteristics of the Hispanic/Latino (H/L) and non-Hispanic White (NHW) pancreatic cancer cohorts.

| Clinical Feature | H/L Cohort<br>n (%) | NHW Cohort<br>n (%) |
| --- | --- | --- |
| **Gender** |  |  |
| Male | 216 (53.1%) | 2032 (52.9%) |
| Female | 191 (46.9%) | 1809 (47.1%) |
| **Tumor Type** |  |  |
| Primary | 407 (100.0%) | 3841 (100.0%) |
| Metastatic | 0 (0.0%) | 0 (0.0%) |
| **Ethnicity** |  |  |
| Spanish/Hispanic | 390 (95.8%) | 0 (0.0%) |
| Spanish NOS; Hispanic NOS; Latino NOS | 11 (2.7%) | 0 (0.0%) |
| Hispanic or Latino | 6 (1.5%) | 0 (0.0%) |
| Non-Spanish; Non-Hispanic | 0 (0.0%) | 3841 (100.0%) |

For statistical analysis, we performed Chi-square tests to evaluate the independence of categorical variables and identify potential associations between ethnicity and pathway-specific alterations. This approach allowed us to determine whether certain molecular disruptions were more prevalent in specific racial/ethnic groups, contributing to a deeper understanding of genomic heterogeneity and potential treatment responses.

To assess overall survival (OS), we employed Kaplan-Meier survival analysis, focusing on the impact of TP53, WNT, PI3K, TGF-Beta, and RTK/RAS pathway alterations. Survival curves were generated to visualize OS probabilities over time, comparing patients based on the presence or absence of these molecular disruptions. The log-rank test was used to identify statistically significant differences between survival curves. Additionally, median survival times were calculated, accompanied by 95% confidence intervals, to ensure the robustness of these estimates. This comprehensive methodological approach provides a nuanced understanding of how pathway-specific genomic alterations may influence prognosis in PC, particularly among H/L patients, and offers critical insights for advancing risk prediction models and precision oncology in diverse populations.

## 3. Results

From the cBioPortal projects that reported ethnicity, we identified and constructed our H/L cohort, which included 407 samples, while the NHW cohort comprised 3,841 samples (Table 1). The gender distribution was similar between cohorts, with the H/L group consisting of 53.1% male and 46.9% female patients, and the NHW group comprising 52.9% male and 47.1% female patients. All patients in both cohorts were diagnosed with primary tumors, with no cases classified as metastatic at the time of data collection. Within the H/L cohort, 95.8% of patients specifically identified as Spanish/Hispanic, while 2.7% were categorized as Spanish NOS, Hispanic NOS, or Latino NOS, and 1.5% identified broadly as Hispanic or Latino. In contrast, all patients in the NHW cohort were classified as Non-Hispanic White, ensuring clear ethnic stratification for comparative analysis.

The comparative analysis of genomic features between H/L and NHW PC patients reveals significant differences (Table 2). The median mutation count was lower in the H/L cohort compared to the NHW cohort, with a highly significant p-value, suggesting potential differences in genomic instability between these populations. However, the median tumor mutational burden (TMB) was similar between both groups, indicating that overall mutation rates may not be a key differentiating factor in tumor biology. In contrast, the median fraction of the genome altered (FGA), a measure of chromosomal instability, showed a trend toward being higher in H/L patients, though not statistically significant. Stratification by Oncotree classification showed that pancreatic adenocarcinoma (PAAD) was the most prevalent diagnosis in both cohorts, with similar distribution across histological subtypes. However, notable differences were observed in specific oncogenic mutations. Within the TGF-Beta pathway, SMAD2 mutations were significantly more prevalent in H/L patients (1.5%, n = 6) compared to NHW patients (0.4%, n = 14) (p = 6.3e-3). Similarly, SMAD4 mutations were detected in 15.0% (n = 61) of H/L patients versus 19.9% (n = 765) of NHW patients (p = 0.02). In the RTK/RAS pathway, mutations in ERBB4 (3.4%, n = 14 vs. 1.8%, n = 70; p = 0.03), ALK (2.7%, n = 11 vs. 1.1%, n = 42; p = 0.01), HRAS (1.2%, n = 5 vs. 0.1%, n = 5; p = 1.0e-4), and RIT1 (0.7%, n = 3 vs. 0.1%, n = 5; p = 0.03) were significantly enriched in the H/L cohort. Furthermore, within the WNT pathway, CTNNB1 mutations were observed in 2.9% (n = 12) of H/L patients compared to 1.3% (n = 51) of NHW patients (p = 0.01). These findings highlight ethnicity-specific molecular variations in PC, suggesting that H/L patients exhibit distinct genomic profiles that may influence tumor progression and response to therapy. Understanding these disparities is crucial for advancing precision medicine strategies tailored to underrepresented populations.

**Table 2.**
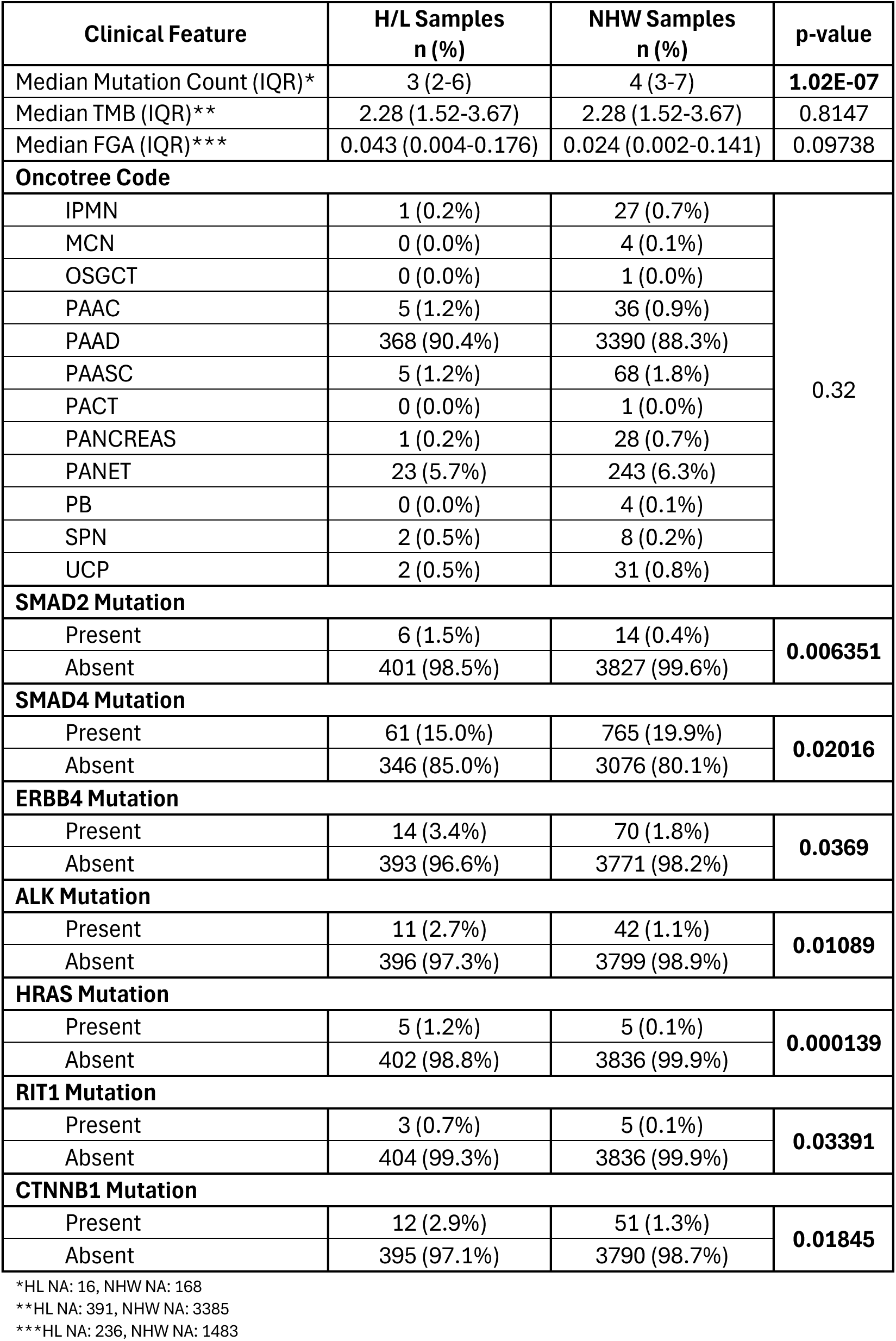
Ethnicity-associated differences in clinical features between Hispanic/Latino (H/L) and non-Hispanic White (NHW) pancreatic cancer cohorts.

| Clinical Feature | H/L Samples<br>n (%) | NHW Samples<br>n (%) | p-value |
| --- | --- | --- | --- |
| Median Mutation Count (IQR)* | 3 (2-6) | 4 (3-7) | **1.02E-07** |
| Median TMB (IQR)** | 2.28 (1.52-3.67) | 2.28 (1.52-3.67) | 0.8147 |
| Median FGA (IQR)*** | 0.043 (0.004-0.176) | 0.024 (0.002-0.141) | 0.09738 |
| Oncotree Code |  |  |  |
| IPMN | 1 (0.2%) | 27 (0.7%) | 0.32 |
| MCN | 0 (0.0%) | 4 (0.1%) |  |
| OSGCT | 0 (0.0%) | 1 (0.0%) |  |
| PAAC | 5 (1.2%) | 36 (0.9%) |  |
| PAAD | 368 (90.4%) | 3390 (88.3%) |  |
| PAASC | 5 (1.2%) | 68 (1.8%) |  |
| PACT | 0 (0.0%) | 1 (0.0%) |  |
| PANCREAS | 1 (0.2%) | 28 (0.7%) |  |
| PANET | 23 (5.7%) | 243 (6.3%) |  |
| PB | 0 (0.0%) | 4 (0.1%) |  |
| SPN | 2 (0.5%) | 8 (0.2%) |  |
| UCP | 2 (0.5%) | 31 (0.8%) |  |
| SMAD2 Mutation |  |  |  |
| Present | 6 (1.5%) | 14 (0.4%) | 0.006351 |
| Absent | 401 (98.5%) | 3827 (99.6%) |  |
| SMAD4 Mutation |  |  |  |
| Present | 61 (15.0%) | 765 (19.9%) | 0.02016 |
| Absent | 346 (85.0%) | 3076 (80.1%) |  |
| ERBB4 Mutation |  |  |  |
| Present | 14 (3.4%) | 70 (1.8%) | 0.0369 |
| Absent | 393 (96.6%) | 3771 (98.2%) |  |
| ALK Mutation |  |  |  |
| Present | 11 (2.7%) | 42 (1.1%) | 0.01089 |
| Absent | 396 (97.3%) | 3799 (98.9%) |  |
| HRAS Mutation |  |  |  |
| Present | 5 (1.2%) | 5 (0.1%) | 0.000139 |
| Absent | 402 (98.8%) | 3836 (99.9%) |  |
| RIT1 Mutation |  |  |  |
| Present | 3 (0.7%) | 5 (0.1%) | 0.03391 |
| Absent | 404 (99.3%) | 3836 (99.9%) |  |
| CTNNB1 Mutation |  |  |  |
| Present | 12 (2.9%) | 51 (1.3%) | 0.01845 |
| Absent | 395 (97.1%) | 3790 (98.7%) |  |

\*HL NA: 16, NHW NA: 168
\*\*HL NA: 391, NHW NA: 3385
\*\*\*HL NA: 236, NHW NA: 1483

In our comparative analysis of pathway-specific genomic alterations in PC among H/L and NHW patients, we identified significant differences in the TGF-Beta pathway, while other pathways exhibited similar alteration frequencies between groups (Table 3). TGF-Beta pathway alterations were significantly less prevalent in H/L patients (18.4%, n = 75) compared to NHW patients (24.4%, n = 937; p = 0.0086), suggesting potential ethnic-specific differences in pathway dysregulation. Conversely, the absence of TGF-Beta pathway alterations was more frequent in the H/L cohort (81.6%, n = 332) compared to NHW patients (75.6%, n = 2904). Alterations in the RTK/RAS pathway were highly prevalent in both groups, with 85.7% (n = 349) of H/L patients and 85.0% (n = 3263) of NHW patients exhibiting mutations (p = 0.722), indicating a shared molecular signature across both populations. Similarly, no significant difference was observed in WNT pathway alterations, which were present in 11.1% (n = 45) of H/L patients and 8.5% (n = 325) of NHW patients (p = 0.1334), suggesting a potential trend that warrants further investigation. PI3K pathway alterations were detected in 11.3% (n = 46) of H/L patients and 11.4% (n = 438) of NHW patients (p = 1), indicating no substantial disparity between the cohorts. TP53 alterations, which are a hallmark of PC, were detected in 69.0% (n = 281) of H/L patients and 70.3% (n = 2700) of NHW patients (p = 0.6396), showing no significant differences between groups. These findings highlight key molecular distinctions in PC, particularly the lower prevalence of TGF-Beta pathway alterations in H/L patients, which may have implications for disease progression, tumor microenvironment interactions, and therapeutic response. Further research is warranted to elucidate the functional impact of these genomic differences and their relevance to ethnicity-specific clinical outcomes.

**Table 3.** Rates of TGF-Beta, RTK/RAS, WNT, PI3K and TP53 pathway alterations among Hispanic/Latino (H/L) and Non-Hispanic White (NHW) pancreatic cancer patients.

|  | H/L Samples<br>n (%) | NHW Samples<br>n (%) | p-value |
| --- | --- | --- | --- |
| TGF-beta Alterations Present | 75 (18.4%) | 937 (24.4%) | **0.008641** |
| TGF-beta Alterations Absent | 332 (81.6%) | 2904 (75.6%) |  |
| RTK/RAS Alterations Present | 349 (85.7%) | 3263 (85.0%) | 0.722 |
| RTK/RAS Alterations Absent | 58 (14.3%) | 578 (15.0%) |  |
| WNT Alterations Present | 45 (11.1%) | 325 (8.5%) | 0.1334 |
| WNT Alterations Absent | 362 (88.9%) | 3416 (88.9%) |  |
| PI3K Alterations Present | 46 (11.3%) | 438 (11.4%) | 1 |
| PI3K Alterations Absent | 361 (88.7%) | 3403 (88.6%) |  |
| TP53 Alterations Present | 281 (69.0%) | 2700 (70.3%) | 0.6396 |
| TP53 Alterations Absent | 126 (31.0%) | 1141 (29.7%) |  |

The Kaplan-Meier survival analysis for H/L PC patients with TGF-Beta pathway alterations demonstrated no statistically significant difference in overall survival between those with and without the alteration (Figure 1). The survival curves closely overlapped, with a p-value of 0.47, suggesting that TGF-Beta pathway mutations do not strongly influence prognosis in this cohort. The shaded confidence intervals highlight variability in survival estimates, indicating potential heterogeneity among patients. However, the relatively small sample size may limit statistical power, necessitating further investigation with larger datasets to clarify the role of TGF-Beta pathway alterations in H/L PC.

**Figure 1.**
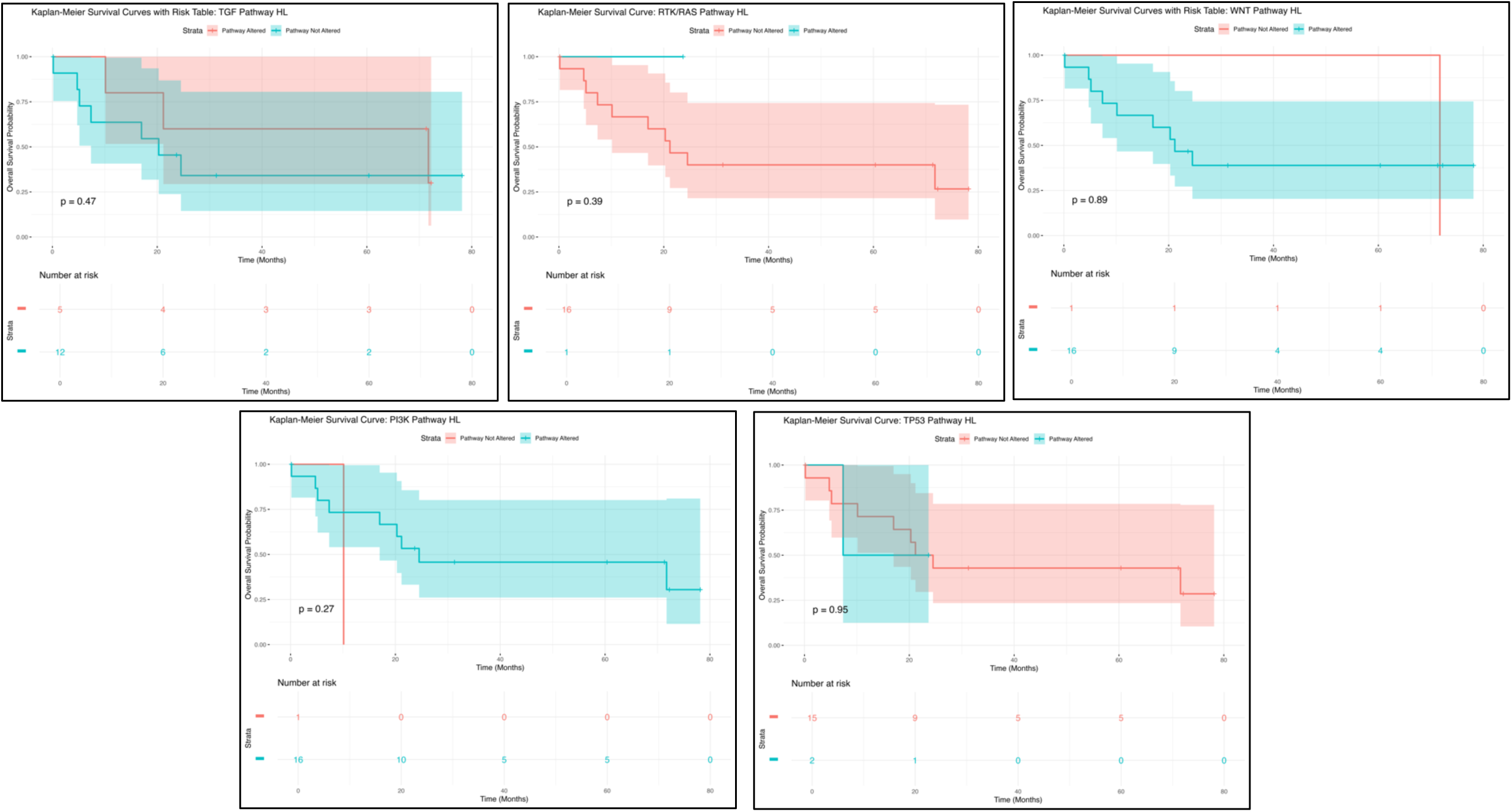
Kaplan-Meier overall survival curves for Hispanic/Latino (H/L) pancreatic cancer (PC) patients, stratified by the presence or absence of TGF-Beta (upper left), RTK/RAS (upper middle), WNT (upper right), PI3K (lower left), and TP53 (lower right) pathway alterations.

For the RTK/RAS pathway, Kaplan-Meier survival analysis also revealed no significant difference in overall survival between patients with and without alterations (p = 0.39) (Figure 1). While the survival trajectory for patients with RTK/RAS pathway mutations exhibited a gradual decline, those without alterations maintained a higher survival probability over time. The broad confidence intervals surrounding the survival curves suggest substantial variability, likely influenced by the limited sample size. These findings indicate that RTK/RAS pathway alterations may not serve as a strong prognostic factor in this cohort, though additional research is warranted to validate these observations.

Similarly, Kaplan-Meier survival analysis for WNT pathway alterations showed no significant association with overall survival in H/L PC patients (p = 0.89) (Figure 1). Patients with WNT pathway alterations displayed a steady decline in survival, while those without alterations had slightly better survival probabilities. However, the broad confidence intervals suggest considerable variability, reinforcing the need for larger studies to assess whether WNT pathway alterations contribute to PC progression or treatment response disparities in this population.

The survival analysis for PI3K pathway alterations in H/L PC patients also showed no statistically significant differences in overall survival (p = 0.37) (Figure 1). The survival curve for the altered group exhibited a gradual decline, whereas patients without PI3K alterations had a more stable trajectory. The wide confidence intervals indicate substantial variation, suggesting that PI3K pathway alterations may not be a strong prognostic marker in this cohort. Further research with expanded sample sizes is necessary to determine whether PI3K pathway mutations influence PC outcomes in H/L patients.

Lastly, the Kaplan-Meier survival analysis for TP53 pathway alterations showed no significant differences in overall survival between H/L PC patients with and without mutations (p = 0.39) (Figure 1). The survival curve for patients with TP53 alterations exhibited a steeper decline early on, while those without alterations showed a more gradual decrease. Overlapping confidence intervals indicate variability in survival estimates, likely due to the small sample size. These findings suggest that TP53 mutations may not have a strong prognostic impact in H/L PC patients, but further validation with larger cohorts is needed to fully elucidate their role in disease progression and therapeutic response.

Overall, these findings highlight the lack of statistically significant survival differences across multiple pathway alterations in H/L PC patients, suggesting that these mutations may not serve as strong prognostic markers in this cohort. The broad confidence intervals and overlapping survival curves indicate substantial variability, likely influenced by the limited sample size. These results underscore the need for larger, more diverse datasets to comprehensively evaluate the molecular drivers of PC in H/L populations and to determine their potential implications for disease progression, therapeutic response, and targeted treatment strategies.

The Kaplan-Meier survival analysis for TGF-Beta pathway alterations in NHW PC patients showed no statistically significant difference in overall survival between those with and without the alteration (Figure S1). The survival curves of both groups were closely aligned, with a p-value of 0.29, indicating that TGF-Beta pathway alterations may not serve as a strong prognostic marker in this cohort. The broad confidence intervals suggest some variability in survival estimates over time, highlighting potential heterogeneity in patient outcomes. While no significant divergence was observed, further studies with larger and more diverse patient populations are necessary to determine the clinical significance of TGF-Beta pathway alterations in NHW PC patients.

In contrast, RTK/RAS pathway alterations in NHW PC patients exhibited a slight trend toward reduced survival compared to those without alterations (Figure S1). The survival curve for RTK/RAS-altered patients (red curve) demonstrated a gradual decline in survival probability relative to the non-altered group (blue curve). However, this difference was not statistically significant (p = 0.61), and the wide confidence intervals suggest high uncertainty, likely influenced by variability in patient outcomes and the limited sample size in the altered group. These findings suggest that RTK/RAS pathway alterations may not have a strong prognostic impact in this cohort, though further analysis with larger cohorts may be needed to explore potential subgroup-specific effects.

The Kaplan-Meier survival analysis for WNT pathway alterations in NHW PC patients also showed no statistically significant association with overall survival (Figure S1). The survival curves for both groups closely overlapped, with the altered group (red curve) and non-altered group (blue curve) following nearly identical trajectories over time (p = 0.65). The broad confidence intervals, particularly at later time points, suggest variability in survival estimates. These findings indicate that WNT pathway alterations may not play a major prognostic role in NHW PC. However, given the biological relevance of WNT signaling in tumor progression, further research using larger datasets and more refined subgroup analyses may help elucidate its clinical impact.

Conversely, the Kaplan-Meier survival analysis for PI3K pathway alterations in NHW PC patients revealed a statistically significant association with poorer overall survival (Figure S1). Patients with PI3K pathway alterations (red curve) demonstrated a more pronounced decline in survival probability compared to those without alterations (blue curve), with a p-value of 0.019, indicating a significant survival disadvantage. The separation between the survival curves suggests that PI3K pathway dysregulation may contribute to disease progression and worse outcomes in NHW PC patients. However, the wide confidence intervals in the altered group highlight variability in survival estimates, potentially due to the limited sample size. These findings emphasize the need for further investigation into the role of PI3K pathway alterations in PC progression and their potential as therapeutic targets.

The Kaplan-Meier survival analysis for TP53 pathway alterations in NHW PC patients similarly revealed a statistically significant association with poorer overall survival (Figure S1). Patients harboring TP53 pathway alterations (red curve) exhibited a marked decline in survival probability compared to those without alterations (blue curve), with a p-value of 0.0472, suggesting a significant survival disadvantage. The separation between the survival curves indicates that TP53 pathway dysregulation may play a crucial role in disease progression and prognosis in NHW PC patients. However, the wide confidence intervals in the altered group suggest some degree of variability in survival estimates, potentially due to the smaller sample size. These findings underscore the importance of further research into the clinical implications of TP53 pathway alterations and their potential role in precision medicine strategies for PC treatment.

Overall, these results highlight pathway-specific differences in survival outcomes among NHW PC patients, with PI3K and TP53 pathway alterations showing significant associations with poorer prognosis, while TGF-Beta, WNT, and RTK/RAS alterations did not appear to strongly influence survival outcomes. The significant impact of PI3K and TP53 dysregulation on survival suggests that these pathways may play a critical role in disease progression and could serve as potential therapeutic targets. These findings underscore the importance of comprehensive genomic profiling in NHW PC patients to better stratify risk and inform targeted treatment strategies. Further studies with larger and more diverse patient populations are needed to refine our understanding of these molecular alterations and their potential integration into precision medicine approaches for PC management.

**Figure S1.** Kaplan-Meier overall survival curves for Non-Hispanic White (NHW) pancreatic cancer patients, stratified by the presence or absence of TGF-Beta (upper left), RTK/RAS (upper middle), WNT (upper right), PI3K (lower left), and TP53 (lower right) pathway alterations.

The analysis of TGF-Beta, RTK/RAS, WNT, PI3K, and TP53 pathway alterations among H/L and NHW PC patients revealed several key differences, indicating potential ethnicity-associated molecular variability (Table S1).

Most genes within the TGF-Beta pathway exhibited low mutation frequencies in both groups, with a few notable differences. SMAD2 mutations were significantly more prevalent in H/L patients (1.5%) compared to NHW patients (0.4%) (p = 0.0064), suggesting potential ethnic variability in this gene’s involvement in PC. Conversely, SMAD4 mutations were more frequent in NHW patients (19.9%) than in H/L patients (15.0%) (p = 0.0202), highlighting a possible disparity in the molecular landscape of this pathway between populations. Other key genes, including SMAD3, TGFBR1, and TGFBR2, showed comparable mutation rates between the two groups, with no statistically significant differences. Notably, ACVR2A and ACVR2B mutations were absent in H/L patients and extremely rare in NHW patients, emphasizing their limited role in PC across both ethnic groups. These findings suggest SMAD2 and SMAD4 as potential key players in ethnicity-associated molecular differences, warranting further investigation into their functional impact on disease progression and therapeutic response.

The RTK/RAS pathway demonstrated notable differences in mutation frequencies between H/L and NHW PC patients (Table S1). ERBB4 mutations were significantly more common in H/L patients (3.4%) than in NHW patients (1.8%) (p = 0.0369), suggesting potential ethnicity-associated differences in this gene’s role in tumor progression. Similarly, ALK mutations were observed at a higher frequency in H/L patients (2.7%) than in NHW patients (1.1%) (p = 0.0109), indicating a possible differential molecular landscape between these populations. While MET mutations were more prevalent in H/L patients (2.0%) compared to NHW patients (0.9%), this difference did not reach statistical significance (p = 0.0671). Other RTK/RAS pathway-related genes, including EGFR, ERBB2, PDGFRA, FGFR family members, IGF1R, RET, and ROS1, exhibited low alteration frequencies with no significant differences between the two groups. These findings suggest that specific RTK/RAS pathway alterations, particularly in ERBB4 and ALK, may be more frequent in H/L PC patients, warranting further research into their functional implications and potential as therapeutic targets.

The WNT pathway analysis revealed distinct mutation patterns between the two groups. CTNNB1 mutations were significantly more common in H/L patients (2.9%) than in NHW patients (1.3%) (p = 0.01845), suggesting potential ethnicity-related variability in WNT pathway dysregulation. Similarly, AXIN1 mutations were more prevalent in H/L patients (1.0%) compared to NHW patients (0.3%), though this difference did not reach statistical significance (p = 0.07244). Other key WNT pathway-related genes, including APC, AXIN2, and RNF43, showed comparable mutation rates between the two groups, with no statistically significant differences observed. APC mutations were detected in 2.9% of H/L patients and 2.4% of NHW patients (p = 0.5803), while RNF43 alterations were slightly more frequent in NHW patients (5.4%) than in H/L patients (3.9%) (p = 0.2392). Notably, several WNT pathway genes, including DKK1, DKK2, DKK3, TCF7, and WIF1, showed no mutations in either group, suggesting an overall low mutational burden in this pathway for PC. These findings suggest that while CTNNB1 and AXIN1 mutations may be more common in H/L patients, additional research is needed to understand their clinical implications in PC progression and treatment response.

The analysis of PI3K pathway alterations revealed low mutation frequencies across both groups, with no statistically significant differences (Table S1). PIK3CA mutations were detected in 1.7% of H/L patients compared to 2.1% of NHW patients (p = 0.7085), while PTEN mutations occurred at similar rates in both groups (1.0% in H/L vs. 1.2% in NHW, p = 1). Other key PI3K pathway genes, including PIK3R1 (0.5% in H/L vs. 0.8% in NHW, p = 0.764), AKT2 (0.5% in H/L vs. 0.3% in NHW, p = 0.3581), and MTOR (2.0% in H/L vs. 1.1% in NHW, p = 0.1466), demonstrated slightly higher alteration frequencies in H/L patients; however, these differences were not statistically significant. Additionally, several PI3K pathway-related genes, such as RHEB, PIK3R2, and PPP2R1A, showed low or absent mutation rates across both cohorts, suggesting a limited mutational burden in this pathway. These findings indicate that PI3K pathway alterations occur at similar rates between H/L and NHW patients, with no clear evidence of ethnicity-specific molecular drivers. Larger studies may be necessary to determine the potential impact of these alterations on PC development and therapeutic response.

The TP53 pathway analysis demonstrated minor differences between H/L and NHW PC patients. TP53 mutations were slightly more prevalent in H/L patients (68.1%) compared to NHW patients (64.7%), though this difference was not statistically significant (p = 0.2015). Similarly, CDKN2A mutations were more frequent in NHW patients (21.0%) compared to H/L patients (17.0%), with borderline significance (p = 0.06467). ATM mutations also showed a trend toward higher prevalence in NHW patients (3.9%) than in H/L patients (2.0%) (p = 0.06441), suggesting possible ethnicity-related differences in DNA damage response mechanisms. In contrast, mutations in other key TP53 pathway genes, including MDM2, MDM4, CHEK2, and RPS6KA3, were rare or absent in both groups, with no statistically significant differences. These findings suggest that while overall TP53 pathway alterations occur at similar rates between H/L and NHW patients, specific mutations in genes like CDKN2A and ATM may reflect subtle ethnic differences that warrant further investigation into their role in PC progression and therapeutic response.

These findings highlight potential ethnicity-specific trends in SMAD2, SMAD4, CTNNB1, ERBB4, and ALK mutations between H/L and NHW PC patients. However, the lack of statistical significance in most comparisons emphasizes the need for larger studies to validate these observations and explore their implications for precision medicine and targeted therapy in H/L PC patients. Understanding these molecular differences could provide critical insights into ethnicity-specific tumor biology and guide more effective treatment strategies for underrepresented populations.

**Table S1.** Alteration rates of TGF-Beta, RTK/RAS, WNT, PI3K, and TP53 pathway-related genes among Hispanic/Latino (H/L) and Non-Hispanic White (NHW) pancreatic cancer (PC) patients.

## 4. Discussion

Pancreatic cancer (PC) remains one of the most lethal malignancies worldwide, with disparities in incidence, disease progression, and survival outcomes across racial and ethnic groups. While NHW patients have historically been the focus of genomic studies, there is a growing recognition of the need to characterize the molecular landscape of PC in H/L populations. Our study sought to address this gap by systematically analyzing alterations in five key oncogenic pathways—TGF-Beta, RTK/RAS, WNT, PI3K, and TP53—among H/L and NHW PC patients. By identifying ethnicity-specific differences in pathway mutations and assessing their prognostic implications, our findings contribute to a more comprehensive understanding of the molecular epidemiology of PC and its potential applications for risk prediction and precision medicine.

### Ethnicity-Specific Molecular Differences in PC

Our results indicate that H/L PC patients exhibit distinct molecular profiles compared to NHW patients, with key differences observed in the TGF-Beta, RTK/RAS, and WNT pathways. In the TGF-Beta pathway, SMAD2 mutations were significantly more common in H/L patients (1.5%) than in NHW patients (0.4%) (p = 0.0064), while SMAD4 mutations were more prevalent in NHW patients (19.9%) than in H/L patients (15.0%) (p = 0.0202). SMAD4 inactivation is associated with poor prognosis and metastasis in PC, and its lower mutation frequency in H/L patients raises the question of whether alternative mechanisms contribute to TGF-Beta signaling dysregulation in this population. The role of epigenetic modifications and tumor microenvironmental factors should be explored in future studies to determine whether SMAD2-driven tumor progression in H/L patients follows a different trajectory than SMAD4-driven pathways in NHW patients.

Within the RTK/RAS pathway, ERBB4 (3.4% vs. 1.8%, p = 0.0369) and ALK (2.7% vs. 1.1%, p = 0.0109) mutations were significantly enriched in H/L patients, suggesting that ethnicity-specific differences in receptor tyrosine kinase (RTK) activation may drive tumorigenesis in this group. While KRAS mutations were similarly frequent in both cohorts (>80%), the increased prevalence of HRAS and RIT1 mutations in H/L patients suggests alternative routes of RAS pathway activation that could have therapeutic relevance. Prior studies have linked HRAS mutations to aggressive tumor behavior, raising the possibility that H/L patients with these alterations may benefit from RAS pathway inhibitors or targeted therapies beyond standard KRAS-directed treatments.

In the WNT pathway, CTNNB1 mutations were significantly more prevalent in H/L patients (2.9% vs. 1.3%, p = 0.01845), while AXIN1 mutations showed a trend toward higher frequency in H/L patients (1.0% vs. 0.3%, p = 0.07244). The WNT/β-catenin signaling pathway is known to contribute to tumor progression, metastasis, and stem cell-like tumor phenotypes, and its increased dysregulation in H/L patients could have clinical implications for tumor aggressiveness and resistance to standard chemotherapy. Given that WNT pathway inhibitors are currently under investigation for multiple cancer types, these findings suggest that H/L patients may represent a distinct molecular subgroup that could benefit from WNT-targeted therapies.

### Clinical Implications for Risk Prediction and Precision Medicine

From a clinical epidemiology and risk stratification perspective, our study highlights the potential for pathway-specific genomic profiling to enhance pancreatic cancer risk prediction models in diverse populations. The increased frequency of SMAD2, ERBB4, ALK, and CTNNB1 mutations in H/L patients suggests that these genes may serve as biomarkers for early detection, prognosis, and therapy selection in this population. Additionally, the differential impact of SMAD4 loss in NHW patients versus SMAD2 alterations in H/L patients raises important questions regarding ethnicity-specific disease progression pathways.

Our Kaplan-Meier survival analysis further underscores the importance of pathway-specific prognostic models. Among NHW PC patients, PI3K and TP53 pathway alterations were significantly associated with worse survival (p = 0.019 and p = 0.0472, respectively), whereas these alterations had no statistically significant survival impact in H/L patients. The poorer prognosis associated with PI3K alterations in NHW patients aligns with previous reports linking PI3K/AKT/mTOR signaling to therapy resistance and tumor progression. Given that PI3K inhibitors are currently in clinical trials, this finding supports ethnicity-informed therapeutic stratification, where NHW patients with PI3K mutations may derive greater benefit from PI3K-targeted agents.

In contrast, the lack of significant survival differences in H/L PC patients across all pathways suggests that tumor biology in this population may be influenced by non-genomic factors. These could include immune microenvironment differences, metabolic reprogramming, or environmental exposures such as high rates of diabetes and obesity. Future studies incorporating multi-omics approaches—including epigenomics, transcriptomics, and metabolomics—will be crucial for unraveling the interplay between genetic and non-genetic determinants of PC progression in H/L patients.

### Limitations and Future Directions

While our study represents one of the most comprehensive ethnicity-specific genomic analyses of PC to date, several limitations must be acknowledged. First, the retrospective nature of our dataset introduces potential selection bias, as publicly available genomic datasets may not fully capture the diversity of the broader PC patient population. Additionally, the relatively small number of H/L patients compared to NHW patients may limit the statistical power of our findings, particularly for low-frequency mutations.

Future studies should aim to validate these findings in larger, prospective cohorts with balanced racial/ethnic representation. Moreover, functional studies are needed to determine whether the observed mutations in SMAD2, ERBB4, ALK, and CTNNB1 lead to distinct tumor phenotypes in H/L patients. The use of patient-derived organoid models (36) and single-cell sequencing (37) could provide critical insights into the biological consequences of these pathway alterations and their impact on tumor heterogeneity, drug resistance, and immune evasion.

Finally, incorporating ethnicity-specific genomic data into clinical decision-making algorithms will be essential for achieving precision oncology in PC. Our findings suggest that standard risk prediction models based solely on NHW patient data may not fully capture the unique molecular drivers of PC in H/L patients, highlighting the urgent need for inclusive and representative genomic studies.

## 5. Conclusions

In summary, our study provides key insights into pathway-specific genomic alterations in pancreatic cancer across diverse racial/ethnic groups. SMAD2, ERBB4, ALK, and CTNNB1 mutations were significantly more frequent in H/L patients, while SMAD4 loss and PI3K pathway alterations were more prognostically relevant in NHW patients. These findings underscore the importance of ethnicity-specific molecular profiling for risk stratification, early detection, and targeted therapy selection. Future efforts should focus on expanding genomic research in underrepresented populations, integrating multi-omics approaches, and developing ethnicity-specific precision medicine strategies to improve pancreatic cancer outcomes for all patients.

## Data Availability

All data used in the present study is publicly available at https://www.cbioportal.org/ and https://genie.cbioportal.org. Additional data can be provided upon reasonable request to the authors.

